# Considerations for using multiple imputation in propensity score-weighted analysis

**DOI:** 10.1101/2021.06.30.21259793

**Authors:** Andreas Halgreen Eiset, Morten Frydenberg

## Abstract

We present our considerations for using multiple imputation to account for missing data in propensity score-weighted analysis with bootstrap percentile confidence interval. We outline the assumptions underlying each of the methods and discuss the methodological and practical implications of our choices and briefly point to alternatives. We made a number of choices *a priori* for example to use logistic regression-based propensity scores to produce “standardized mortality ratio”-weights and Substantive Model Compatible-Full Conditional Specification to multiply impute missing data (given no violation of underlying assumptions). We present a methodology to combine these methods by choosing the propensity score model based on covariate balance, using this model as the substantive model in the multiple imputation, producing and averaging the point estimates from each multiple imputed data set to give the estimate of association and computing the percentile confidence interval by bootstrapping. The described methodology is demanding in both work-load and in computational time, however, we do not consider the prior a draw-back: it makes some of the underlying assumptions explicit and the latter may be a nuisance that will diminish with faster computers and better implementations.

## Introduction

In this paper we present the considerations behind estimating the change in prevalence of post-traumatic stress disorder (PTSD) associated with long-distance migration using multiple imputation to handle missing data, propensity score-weighting to adjust for confounding and bootstrap to produce a percentile confidence interval. We will focus on the many statistical methodological problems we encountered and refer the reader to the accompanying paper (1) for the subject matter problem. The relevant data consisted of a 20-items questionnaire and a clinical examination including assessment of possible psychiatric disorders, applied to a sample of Syrian asylum seekers in Denmark and a sample of Syrian refugees in Lebanon. The outcome, PTSD, was assessed using the “Harvard Trauma Questionnaire” part IV (2), giving a score from 1 to 4 with 2.5 being the commonly used cut-off-score for PTSD.

In a propensity score-weighted analysis you first estimate the propensity given a relevant set of predictors, *Pr* (*E*=1∣*Z*), for each individual in the study population, *ê*_*i*_. The association between long-distance migration and PTSD was estimated as the prevalence among those who migrated to Denmark minus a weighted average of the prevalence of PTSD among refugees who migrated to Lebanon, using weights equal to *ê*_*i*_ / (1*-ê*_*i*_). This requires a number of decisions including: Which covariates to include in the propensity score model? What complexity of the model to use? How to deal with extreme weights? And how to calculate the standard error of the parameter of interest? As we had missing data in the covariates and PTSD status, we set out to combine the propensity score-weighted analysis with multiple imputation. This raised additional questions such as: What are the required assumptions of the missing data process? What is the substantial model and what variables should be included in the model? How to combine the multiple imputations with the propensity score analysis? How to find a valid confidence interval for the parameter of interest? In the following sub-sections we outline the problems we had to consider and the underlying theory. In Methods we discuss our considerations on how to implement these in our specific study and in Results we provide details on our final implementation. The problems, theory, considerations and our decisions are summarized in Table 1, 2 and 3.

**Table 1:** Considerations and decision for building the propensity score model.

| Problem | Theory | Considerations | Decision |
| --- | --- | --- | --- |
| What covariates should be included in the model? | Confounders and potential confounders must be included in the propensity score model | Subject matter knowledge and thorough discussions in the group of authors were undertaken including drawing the assumed association in a directed acyclic graph. | Data were collected on variables of interest including age, sex, mental health status, exposure to violence, socioeconomic status. |
| What complexity of the model to use in the analysis? | Increasing model complexity should be examined to choose the model that obtain covariate "balance" between exposure groups. This is generally assessed subjectively. There are some consensus that balance is obtained when the standardized absolute mean difference are $< 0.10$ for all covariates. | Three models of increasing complexity was defined *a priori*. A threshold of 0.10 was used to define balance of covariates between exposure groups. For each combination of model complexity the missing data was imputed once and covariate balance was plotted. The least complex model with the least truncation that obtained balance were chosen as the propensity score model in the analysis. | The propensity score model with no interaction terms (i.e. the "simple" model). Because of numerical issues two levels of two categorical variables were collapsed. |
| How to handle extreme weights? | For example truncation, i.e. forcing extreme weights to a given threshold, shrinkage, i.e. "pushing" all weights towards the mean altering extreme weights relatively more than weights closer to the mean, or penalization, i.e. constraining the coefficients of the propensity score model which will result in less extreme predicted chance of exposure and thus less extreme weights. | To lower the complexity of the methodology we focused on truncation. Sets of truncation percentiles at 0 & 100, 1 & 99, and 5 & 95 were examined for each of the three complexities of the model | Truncation at 1st and 99th percentiles. |

**Table 2:**
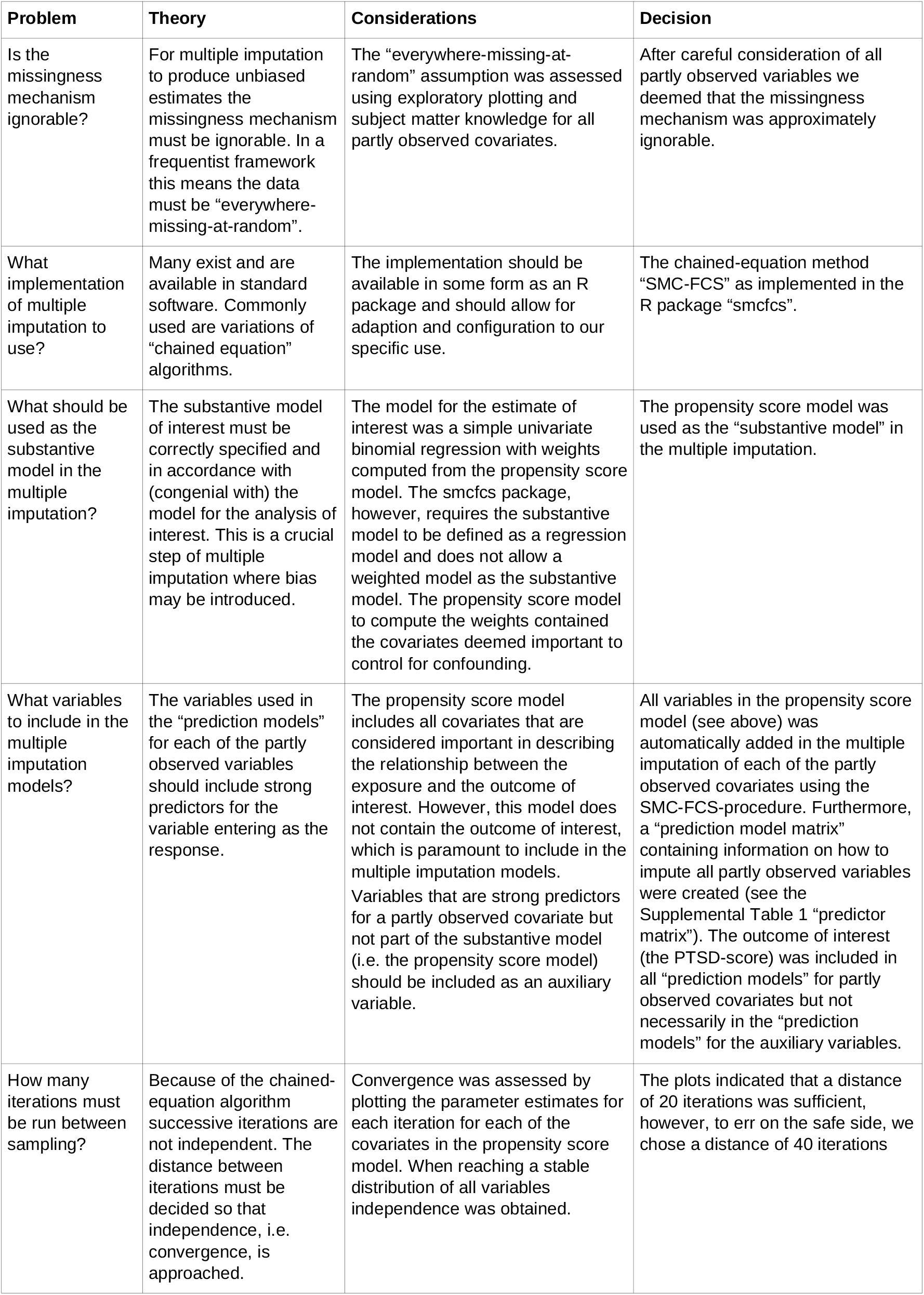
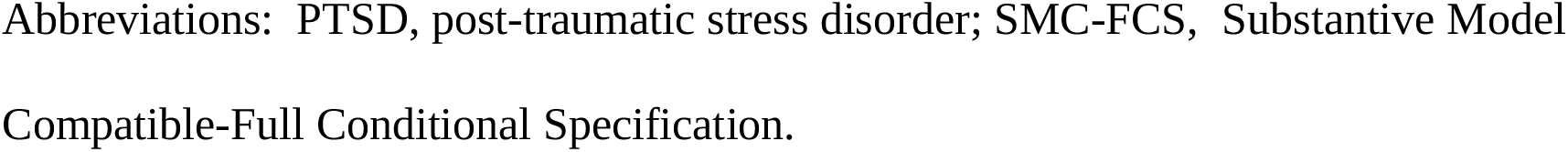
Considerations and decision for building the multiple imputation model.

| Problem | Theory | Considerations | Decision |
| --- | --- | --- | --- |
| Is the missingness mechanism ignorable? | For multiple imputation to produce unbiased estimates the missingness mechanism must be ignorable. In a frequentist framework this means the data must be “everywhere-missing-at-random”. | The “everywhere-missing-at-random” assumption was assessed using exploratory plotting and subject matter knowledge for all partly observed covariates. | After careful consideration of all partly observed variables we deemed that the missingness mechanism was approximately ignorable. |
| What implementation of multiple imputation to use? | Many exist and are available in standard software. Commonly used are variations of “chained equation” algorithms. | The implementation should be available in some form as an R package and should allow for adaption and configuration to our specific use. | The chained-equation method “SMC-FCS” as implemented in the R package “smcfcs”. |
| What should be used as the substantive model in the multiple imputation? | The substantive model of interest must be correctly specified and in accordance with (congenial with) the model for the analysis of interest. This is a crucial step of multiple imputation where bias may be introduced. | The model for the estimate of interest was a simple univariate binomial regression with weights computed from the propensity score model. The smcfcs package, however, requires the substantive model to be defined as a regression model and does not allow a weighted model as the substantive model. The propensity score model to compute the weights contained the covariates deemed important to control for confounding. | The propensity score model was used as the “substantive model” in the multiple imputation. |
| What variables to include in the multiple imputation models? | The variables used in the “prediction models” for each of the partly observed variables should include strong predictors for the variable entering as the response. | The propensity score model includes all covariates that are considered important in describing the relationship between the exposure and the outcome of interest. However, this model does not contain the outcome of interest, which is paramount to include in the multiple imputation models. Variables that are strong predictors for a partly observed covariate but not part of the substantive model (i.e. the propensity score model) should be included as an auxiliary variable. | All variables in the propensity score model (see above) was automatically added in the multiple imputation of each of the partly observed covariates using the SMC-FCS-procedure. Furthermore, a “prediction model matrix” containing information on how to impute all partly observed variables were created (see the Supplemental Table 1 “predictor matrix”). The outcome of interest (the PTSD-score) was included in all “prediction models” for partly observed covariates but not necessarily in the “prediction models” for the auxiliary variables. |
| How many iterations must be run between sampling? | Because of the chained-equation algorithm successive iterations are not independent. The distance between iterations must be decided so that independence, i.e. convergence, is approached. | Convergence was assessed by plotting the parameter estimates for each iteration for each of the covariates in the propensity score model. When reaching a stable distribution of all variables independence was obtained. | The plots indicated that a distance of 20 iterations was sufficient, however, to err on the safe side, we chose a distance of 40 iterations |

Abbreviations: PTSD, post-traumatic stress disorder; SMC-FCS, Substantive Model
Compatible-Full Conditional Specification.

**Table 3:** Consideration and decision for combining multiple imputation and propensity score-weighting and obtaining valid confidence interval

|  | Problem | Theory | Considerations | Decision |
| --- | --- | --- | --- | --- |
| Combining multiple imputation and propensity score-weighted analysis | What sequence of multiple imputation and propensity score-weighted analysis should be implemented? | The “within” procedure (impute the missing data, compute the propensity score-weights and the estimate of association, combine by taking the average to produce the estimate of association) has been proposed as less prone to introduce bias compared with the “across” procedure (impute the missing data, compute the propensity score, combine the propensity scores by taking the average, compute the estimate of association). |  | The “within” procedure was implemented. |
| Obtaining confidence interval | How to account for the uncertainty introduced in both the multiple imputation and propensity score estimation? | Rubin’s rules with modification to large-sample variance estimator or bootstrap has been proposed. | There are some theoretical evidence that bootstrap produces valid estimates of uncertainty in implementations such as the one we propose, however, it has received little attention in applied epidemiology. | Bootstrapping of the “within” procedure was decided upon. |
| Bootstrapping | What type of bootstrap confidence interval and how many bootstraps to produce the confidence interval? | Several types of bootstrap confidence intervals have been proposed, among others the normal, percentile and BCa. For the prior a relatively low number of bootstraps are sufficient, however, it relies on the normal distribution of the estimates. The latter requires a much larger number of repeats, often at least 1000 and are thus computationally intensive. | The percentile type requires less repetitions than the BCa and relaxes the distributional assumption of the normal bootstrap somewhat, however, may still be severely biased in a skewed distribution. | The percentile method with 999 bootstrap samples was used. To assess the influence of the bootstrap confidence interval type on the interpretation of the result we produced four different confidence intervals (normal, basic, percentile and BCa). |
|  | How many data sets should be imputed for each bootstrap? | Since we are using bootstrap to calculate the confidence interval the number of imputations for each missing data point can be kept to a minimum, some recommending as little as two. | We had a relatively high proportion of missing in several variables and for several observations. | The number of imputations was chosen to 10. |

### The propensity score analysis

Table 1 provides an overview of the considerations and decision for building the propensity score model. The relevant predictors to include in the propensity score model are covariates that (potentially) confound the relationship between the exposure and the outcome. The outcome itself and variables that are only associated with the exposure should not be included in the model (3). The complexity of the regression model should be examined so that balance is obtained for all covariates between exposure groups. Extreme weights may lead to suboptimal covariate balance and unstable estimates and are most often remedied by smoothing or truncation, at the cost of potentially introducing bias (4). The estimate of association When only considering the propensity score-weighted analysis, the confidence intervals can be produced by applying some approximate formula to obtain a standard error or via bootstrapping.

### Missing data

The statistical properties of many missing data methods rely on the hypothesized missingness mechanism. The primary interest in applied epidemiology, is whether the missing data mechanism is ignorable, that is, if valid inference can be drawn despite of missing data. In many applied papers using multiple imputations (MI) the authors states that the data is “missing at random” (MAR) and “as a consequence” the inference based on MI is valid. We briefly consider the definition and importance of “missing data” drawing primarily on Seaman *et al*., (5). Very loosely speaking, data is MAR, if the risk of a data point being missing does not depend on the unobserved values, but only on the observed values. However, this is only a superficial definition. The terminology “missing at random” (MAR) and “missing completely at random” (MCAR, which imply MAR) has been in use at least since Rubin’s 1976 paper (6) and was recently extended to include “realized” and “everywhere” versions of both MAR and MCAR (5). In the latter paper the definition is based on parametric models for both the data, *Z*, (which include both outcome variable, *Y*, and covariates, *X*) and the missingness indicator vector, *M*, (which for each entry in *z*, specify if it is observed). Note, we do not observe the entire *z*, but only the entries, where the corresponding entry in m is 1 and we let *o*(*z, m*) denote the observed part of the data, *z*. Furthermore we let *f*_*θ*_ (*z*) denote the density for the data and *Pr*_*φ*_ (*m*|*z*) the conditional probability of the missing pattern, *m*, given the data *z*, with the parameters (*φ, θ*) ∈*Ω*. In a specific study we have the realized data 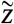 and missing indicator vector 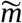 with the realized observed data 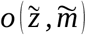.

Example 1. Consider a very small data set with four refugees and four variables: “year of residency”, “sex”, “host country” and “PTSD-status”. One realization could be:

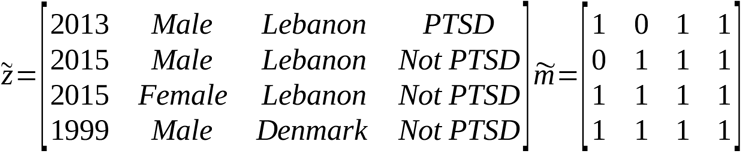

With the realised observed data 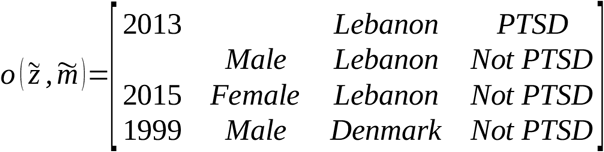

Data is said to be realized-MAR if for all *φ*, 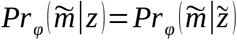 for all *z*, where 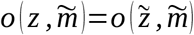 that is, the probability of the realized missingness pattern 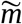 is the same for all data *z* that has an observed part that is identical to the realized observed data, i.e. the unobserved part is of no interest. In Example 1, the data is realized-MAR, if the conditional probability of data on “sex” for observation number 1 and data on “year of residency” for observation number 2 are missing and all other entries are observed, does not depend on the value of the missing sex and year of residency as long as all the observed entry is as realized. This is a statement only focusing on the realized missingness pattern and the realized observed data; we do not consider other possible missingness patterns or other possible realizations of the data. To emphasize: it is irrelevant whether for instance “sex” on observation number 2 or “country” on observation number 3 could be missing,

The data generating process is said to be everywhere-MAR if for all *φ* and all *m, Pr*_*φ*_ (*m*|*z*)=*Pr*_*φ*_ (*m*|*z’*) for all *z* and *z’*, where *o*(*z, m*)=*o*(*z’, m*). That is, data is every-where-MAR if it is realized-MAR for all possible realizations and not only for the actu ally observed realization of the missingness pattern and data. Returning to Example 1, when assuming everywhere-MAR the realized data set is irrelevant: We have to check the whole set of possibly missing data conditional probabilities, *Pr*_*φ*_ (*m*|*z*) for all param-eter values, *φ*.

The elaborations above was necessary to qualify the question of interest: Is the missingness mechanism ignorable? That is, when can we make valid inference about the parameter of interest, *θ*, only based on the observed data? Seaman *et al*. (5), illustrated that the answer depends on the type of statistical inference framework and in the “frequentist likelihood framework” you need the missingness mechanism to be everywhere-MAR (and the parameters (*φ, θ*) be variation independent, i.e *Ω*=*Ω*_*φ*_ *×Ω*_*θ*_). So in order to ig-nore the missingness mechanism we have to argue that it is reasonable to assume every-where-MAR. This implies that, for all possible missingness patterns and corresponding observed data, it is reasonable to assume that the risk of that specific pattern does not depend on the value of the missing data but only of the observed data. This is of course an impossible task without some insight into why data is missing in the study. One way to start off is to assume that the missing data mechanism is identical and work indepen-dently from person to person, which reduce the problem to a discussion of the mechanism for a single person.

For example, in Example 1, we have no missing in “PTSD” in the realized observed data, however, we can easily imagine this information missing in another realization of the study. If we assume identical and independent missing mechanism, we have to think of why “year of residency”, “sex”, and “PTSD” could be missing for a person and if the risk of this is independent of the unobserved values given what we have observed for that person. For example, if we only observe “country”, we have to argue, that the risk of this is the same for all individuals in each country, i.e. it does not depend on year, sex or whether or not the person has PTSD. We note that the assumption of independent missingness mechanism might easily be invalid, for example missingness could depend on some unobserved event common for several persons in the study. In the accompanying paper (1) we discuss the everywhere-MAR assumption in the specific study.

In the following, we will assume that the purpose of the data analysis is to estimate *β*, typically a vector of regression coefficients based on the proposed model for the analysis of interest—i.e. the substantive model—of *Y* given the covariates *X*: *Pr* (*y*∣*x; β*).

### Multiple imputations

Table 2 gives an overview of the considerations and decision for using multiple imputation to deal with missing data. Many statistical methods assume no missing data or missingness mechanism MCAR and will produce biased estimates otherwise (7). A popular way to deal with missing data is to use multiple imputation which gives unbi-ased estimates assuming ignorable missingness mechanism and correctly specified multiple imputation model (8,9). Briefly, multiple imputation consists of producing a number, *K*, of data sets with imputed values for the missing data and analyze these complete data sets as planned, resulting in *K* estimates of *β* which are combined, typically by taking taking the average, into a final estimate for *β*. When implemented, the imputation is done for each variable with missing data (a) specifying a regression model for the conditional distribution of the variable given the other (relevant) variables (b) using the observed data to estimate the parameters in this model (c) impute the missing values of the variable by simulating from the Bayesian posterior predictive distribution. The last two steps will in general be taken care of by a software program, as long as the imputation regression models are chosen within the most common regression model families. Often one or several of the “predictor” variables in the imputation regression will have missing values too, resulting in a so-called “chained equation”, that is, the imputed values in one variable are needed to impute the values in another variable and vice versa. Luckily, many software packages can solve this problem using iterative methods. Thus, after deciding on what implementation of multiple imputation to use we are left with problem (a): How to specify the imputation regression models, i.e., what should be used as the substantive model in the multiple imputation, what variables to include in the multiple imputation models and how many iterations must be run between sampling? It has been known for a while that you can introduce bias in the estimation of *β*, if you do not take care in this specification (10). This can happen if the relationship between *y* and *x* in the substantive model is more complicated than the relationship between *x* and *y* in the implemented imputation regression models. For example, if you do not include *y* in the imputation regression model for the covariate *x*_*i*_ then the imputed data for *x*_*i*_ will be unrelated with *y* and, as a consequence, you will underestimate the regression coefficient *β*_*i*_ relating *y* to *x*_*i*_ in the substantive model. Furthermore, if *x*_*i*_ and *x*_*j*_ interact in the substantive model for *y*, then *y* and *x*_*j*_ should (at least) interact in the imputation model for *x*_*i*_ to avoid bias in the estimate of the magnitude of the interaction. It is difficult, even for relatively simple substantive models, to determine how to specify the imputation models in order to avoid this problem. Luckily there exist a statistical method that can combine a specification of the substantive regression model, *y* on *x*, with univariate regression models for each of the variables in *x* given the rest of the *x*’s, into an imputation algorithm (11). This Substantive Model Compatible-Full Conditional Specification (SMC-FCS) algorithm has been implemented in R and Stata for a set of standard regression models (12,13). As the SMC-FCS algorithm is an iterative algorithm, it will not generate independent samples. This implies that one cannot use subsequent samples but only use samples with a specific interval between them.

### Bootstrapping

Table 3 gives an overview of the considerations and decision combining propensity score-weighting and multiple imputation and obtaining a valid confidence interval. Non-parametric bootstrapping is a method to find an approximate confidence interval for a parameter, when applying a specific estimation algorithm to a data set. In bootstrapping the only input is the data set and the estimation algorithm and no assumption is made concerning the distribution or the estimation algorithm. However, the realized sample is assumed to be independent and representative of the target population (14). In the simple bootstrap, the estimation algorithm is applied to the original data and to a number of bootstrap samples, i.e. artificial data sets with the same number of observations as the original, but with the observations being sampled randomly with replacement from the original data set. This results in the original estimate and a set of bootstrap estimates from which a 95% confidence interval can be produced as (a) the original estimate +/- 1.96 times the standard deviation of the bootstrap estimates or (b) the 2.5th and 97.5th percentile of the bootstrap estimates. The first strategy will typical require a relative small number of bootstrap samples, but rely on approximate normality of the estimates, while the second require a large number of bootstrap samples, but does not require any assumptions about the distribution of the estimates.

## Methods

Based on the theoretical considerations above we outline our estimation algorithm. The analysis plan was defined *a priori* and included a number of decisions:

1. the exposure (long-distance migration), outcome (PTSD) and potential confounders (age, sex, socioeconomic status, experienced trauma and mental well-being) (see also (1))
2. addressing of confounding by logistic regression-based propensity score modeling and of missing data by multiple imputation
3. three propensity score models of increasing complexity were defined and three levels of weight truncation (no truncation, truncating at the 1st and 99th percentile, or truncating at the 5th and 95th percentile) were examined for covariate balance (15,16). Based on a single imputed data set for each of the three complexities of the propensity score model, the least complex model with the least amount of truncation to obtain acceptable balance, defined as the absolute standardized difference of ≤ 0.10 on all covariates (15) was chosen for the analysis. See supplementary materials and Figures in (1) for details on the specific models and the exploratory plots
4. given ignorable missingness mechanism, the missing data were multiple imputed using the SMC-FCS algorithm with the chosen propensity score model as the substantive model
5. for each of the multiply imputed data sets: the propensity scores were computed using the chosen propensity score model, converted into weights and the weighted point estimates produced
6. the mean of the point estimates from 5) was the estimate of interest
7. the 95 percentile confidence interval was produced by bootstrapping steps 4-6 a large number of times.

It should be noted that the existing implementation of the SMC-FCS algorithm does not cover our substantive model, the propensity score-weighted analysis, consequently, we decided to use the model for the propensity score as our substantive model. For each partially observed covariate we specified a “prediction model” meaning a regression model to predict the missing value of a partly observed covariate (the response in the regression model in question) given the PTSD score and any additional covariates as deemed relevant based on subject matter insight and exploratory plots. When entering as the response variable, all continuous partially observed covariates were modeled using linear regression with relevant transformation and all discrete covariates were modeled using logistic, multinomial or proportional odds regression. When entering as “predictor variables”, all continuous covariates were modeled as restricted cubic splines with knots at the 10th, 50th and 90th percentiles; all discrete covariates and interactions entered unaltered (see supplementary materials in (1)). The sampling interval between the imputations was decided based on plots of the parameter estimates against the sampling interval.

To combine propensity score-weighting and multiple imputation to produce the estimate of association we used the “within” procedure (17,18): a number of data sets were imputed, for each data set the prevalence difference of PTSD according to long-distance migration was estimated and averaged to give the point estimate (“impute, compute, combine”). The 95-percentile confidence interval was found by bootstrapping this procedure. The procedure is illustrated in Figure 1.

**Figure 1:**
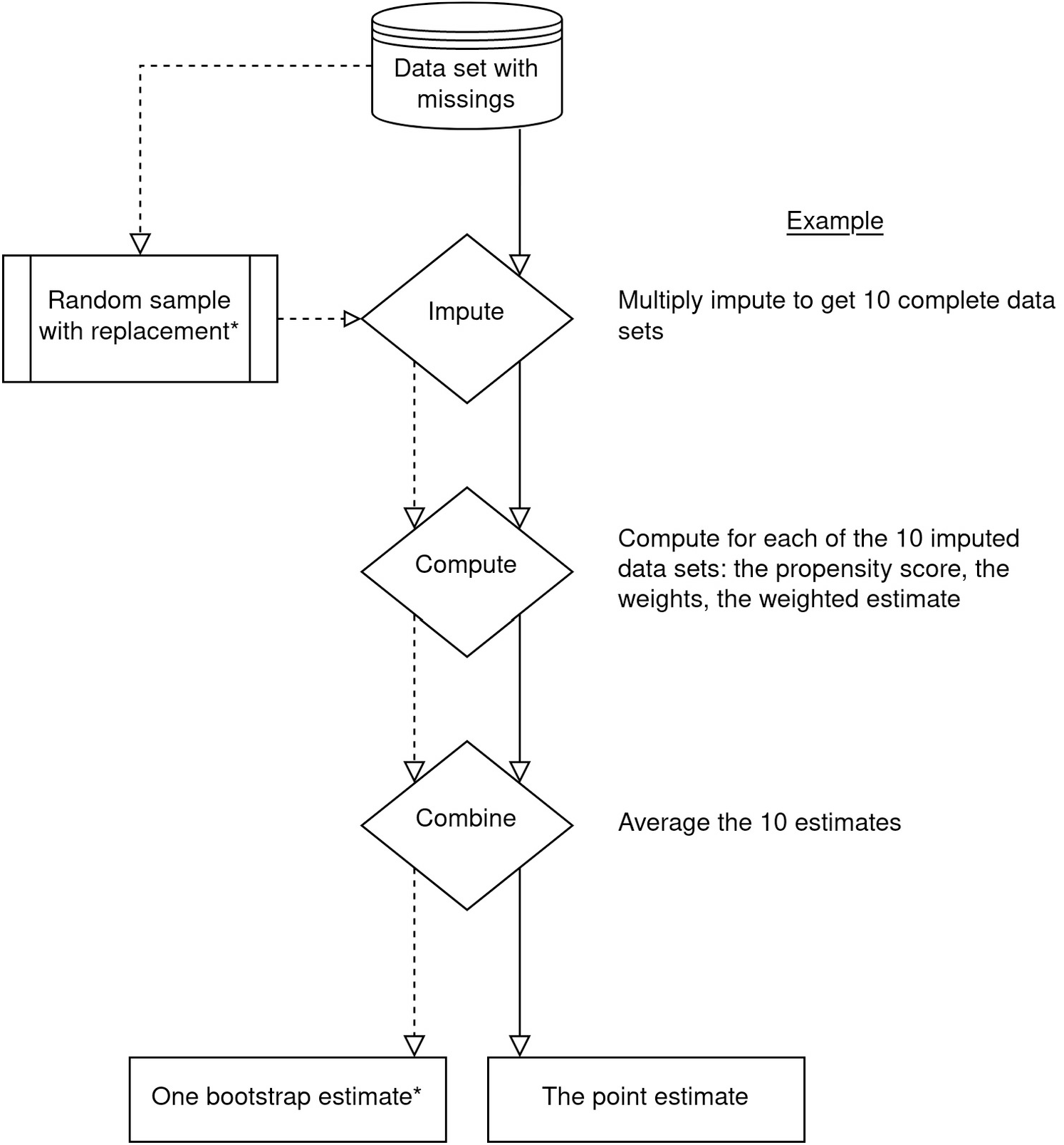
Flow-chart of proposed methodology to combine multiple imputation and propensity score weighting. *The bootstrap is repeated multiple times, for example 1000, to be able to estimate the 95-percentile confidence interval.

All data management, analysis and plots were done in R (19) with heavy reliance on packages “smcfcs” (12) for SMC-FCS multiple imputation; “WeightIt” (20) and “cobalt” (21) for estimation of propensity score weights and assessment of covariate balance; “boot” (22) for parallelized bootstrapping; “furrr” (23) for further parallelizing procedures; and “tidyverse” packages (24) for data wrangling and plotting. The code was run on two Ubuntu systems (18.04.5 and 20.04.1) and a Windows 10 system; all running R 4.0.3. The analysis plan and all R code for analysis and plots, including the specific settings in each procedure are available from https://github.com/eiset/ARCH.

## Results

The simple propensity score model (no interaction terms) with weight truncation at 1st and 99th percentile obtained acceptable balance on all covariates and was chosen as our model. Unfortunately, but not surprisingly, we had to modify our first choice of substantive model (i.e. the propensity score model used in the imputation) due to computational/numerical problems by collapsing two levels of one of the substantive model covariates and two levels of one of the auxiliary covariate (1).

This slightly modified propensity score model was the substantive model in the SMC-FCS multiple imputation and regression models were set up for all partially observed covariates: For example, for imputing the continuous covariate “Age”, the logarithmic transformation of Age, “log Age”, was modeled with covariates from the substantive model entering as predictor variables: “Socioeconomic status”, “PTSD” (as restricted cubic spline) and auxiliary regressors: “Highest education”, “Number of children”, “Systolic blood pressure” (as restricted cubic spline), and “Marital status”. The Age variable was then passively imputed from “log Age” by exponentiating. All partially observed auxiliary variables were also imputed. The “predictor matrix” in Supplemental Table 1 gives details on models for all partly observed variables.

We set the number of imputations to 10 which is well beyond what is often considered sufficient (25). The convergence plots showed that a sampling interval between imputations, i.e. iterations, of 20 was sufficient; to err on the safe side, we chose 40 iterations. Following recommendations of Carpenter and Bithell (14) we produced 999 bootstrap estimates to compute the 95-percentile confidence interval. For practical reasons, three different computers were used to run the final analysis. The time to run 250 bootstrap estimates was from two to 10 hours depending on the system.

The analysis showed an increased prevalence amounting to 8.76 percentage points (95-percentile confidence interval [-1.39; 18.62 percentage points] with little variation in the sensitivity analysis. We refer to the accompanying paper for discussions of the results (1).

## Discussion

In this paper we describe the statistical methodological considerations for combining propensity score-weighting and multiple imputation of missing data. We discuss the assumptions behind both propensity score-weighted estimation and multiple imputation.

In our approach, the substantive model of interest and covariates to include in the propensity score model was explicit. It has been suggested that machine learning or “black box” algorithms may provide reasonable propensity score-weights (4,26,27), however, at the cost of control over the substantive model which is paramount in fulfilling one of the assumptions of multiple imputation: a correctly specified substantive model of interest. And as Bartlett *et al*. notes “We do not consider the requirement to specify a substantive model at the imputation stage to be a shortcoming…” (11). We truncated extreme weights as advocated by several (4,16,28), acknowledging that the decrease in variance comes at the cost of possibly introducing bias. Stabilized weights is another approach to decrease the variance but comes at a similar cost (29); a recent paper (28) found that when estimating the hazard rate by propensity score-weighted Cox regression the choice between ordinary propensity score-weighting (in this case using weights to produce the “average treatment effect”) or its stabilized version made no difference on the confidence interval coverage and that bootstrap gave the least biased variance estimates with best confidence interval coverage.

The SMC-FCS algorithm (11) allows defining the substantive model of interest and imputation models for each partially observed variable and takes care of combining these in the multiple univariate imputations. This may increase the possibility to define a correctly specified multiple imputation model. While model misspecification is considered the overarching source to bias in propensity score modeling (4,30) recent studies suggest that misspecification of the multiple imputation model may not be detrimental in obtaining valid percentile confidence interval when applying a methodology as proposed in this paper (31). We have based our propensity score model on the available evidence and subject matter knowledge, however, recognize the possibility of some remaining bias, for example from residual confounding and from the collapsing of two levels of one of the substantive model variables. Seaman and White (32) showed that the “within” procedure as proposed by Qu and Lipkovich (33) gives unbiased point estimates assuming ignorable missingness mechanism and that including a “missing-value indicator variable” in the data set may reduce bias when the missingness mechanism is not ignorable, however, increase bias when the it is ignorable. We subjected every variable to careful examination and are satisfied that the “everywhere-MAR” assumption is not violated, however, we acknowledge that this is subject to discussion and cannot be guaranteed. We used bootstrap to produce a 95-percentile confidence interval taking into account uncertainty introduced by modeling in both the propensity score and multiple imputation step. Alternatively, the “Rubin’s rule” are used in several studies and are the traditional choice when doing multiple imputation (without propensity score modeling). Qu and Lipkovich (33) noted that “Rubin’s rule” does not account for the uncertainty introduced in the propensity score estimation and, thus, is not valid in theory while others note that it may produce valid estimates in practice (32). There is no clear evidence on what step to bootstrap when combining propensity score-weighting and multiple imputation (34). In our approach, we bootstrapped the entire “within” procedure to produce a confidence interval that accounts for all uncertainty introduced by modeling. This procedure is similar to that applied to a simple simulated data set with ignorable missingness mechanism by Penning de Vries and Groenwold (18). Schomaker and Heumann (34) suggest that bootstrapping after multiply imputing the data sets may produce similar results at lower computational expense, however, a later study (31) found that this may increase bias compared with bootstrapping the entire procedure.

Our proposed methodology takes several hours to run on “standard” laptop computers and we experienced numerical problems with strata with relatively few observations. Going forward, we are eager to examine the sensitivity of our result to different methodologies for example using other *g-*methods such as *g-*computation or other multiple imputation methods such as machine learning algorithms. The produced point estimate and confidence interval could also be compared to alternative methods that lowers the computing time such as “Rubin’s rules” or the recently proposed “von Hippel” method for using bootstrap in multiple imputation (though does not include propensity score modeling) (35).

In this article we have striven to make clear the many choices that we had to go through to produce the estimate of interest. It is our hope that others can make use of our experience in planning their research, creating the analysis plan and running their analysis.

## Data Availability

Not applicable

## Notes

### Competing Interest Statement

The authors have declared no competing interest.

### Funding Statement

No external funding was received for this work

## References

1. Eiset AH, Aoun MP, Stougaard M, et al. The association between post-traumatic stress disorder and long-distance migration: A cross-sectional study of refugee health. In review

2. Mollica RF, Caspi-Yavin Y, Bollini P, et al. The Harvard Trauma Questionnaire: Validating a Cross-Cultural Instrument for Measuring Torture, Trauma, and Posttraumatic Stress Disorder in Indochinese Refugees. J. Nerv. 1992;180(2):111–116.

3. Brookhart MA, Schneeweiss S, Rothman KJ, et al. Variable Selection for Propensity Score Models. Am. J. Epidemiol. 2006;163(12):1149–1156.

4. Lee BK, Lessler J, Stuart EA. Weight Trimming and Propensity Score Weighting. PLOS ONE. 2011;6(3):e18174.

5. Seaman SR, Galati J, Jackson D, et al. What Is Meant by “Missing at Random”? Stat. Sci. 2013;28(2):257–268.

6. Rubin DB. Inference and Missing Data. Biometrika. 1976;63(3):581–592.

7. Little RJA. Regression With Missing X’s: A Review. J. Am. Stat. Assoc. 1992;87(420):1227–1237.

8. Rubin DB. Multiple Imputation After 18+ Years. J. Am. Stat. Assoc. 1996;91(434):473–489.

9. Sterne JAC, White IR, Carlin JB, et al. Multiple imputation for missing data in epidemiological and clinical research: potential and pitfalls. BMJ [electronic article]. 2009;338. (http://www.ncbi.nlm.nih.gov/pmc/articles/PMC2714692/). (Accessed June 24, 2015)

10. Seaman SR, Bartlett JW, White IR. Multiple imputation of missing covariates with non-linear effects and interactions: an evaluation of statistical methods. BMC Med. Res. Methodol. 2012;12:46.

11. Bartlett JW, Seaman SR, White IR, et al. Multiple imputation of covariates by fully conditional specification: Accommodating the substantive model. Stat. Methods Med. Res. 2015;24(4):462–487.

12. Bartlett JW, Keogh R. smcfcs: Multiple Imputation of Covariates by Substantive Model Compatible Fully Conditional Specification. 2020.(https://CRAN.R-project.org/package=smcfcs)

13. Bartlett JW, Morris TP. Multiple Imputation of Covariates by Substantive-model Compatible Fully Conditional Specification. Stata J. Promot. Commun. Stat. Stata. 2015;15(2):437–456.

14. Carpenter J, Bithell J. Bootstrap confidence intervals: when, which, what? A practical guide for medical statisticians. Stat. Med. 2000;19(9):1141–1164.

15. Austin PC. Balance diagnostics for comparing the distribution of baseline covariates between treatment groups in propensity-score matched samples. Stat. Med. 2009;28(25):3083–3107.

16. Austin PC, Stuart EA. Moving towards best practice when using inverse probability of treatment weighting (IPTW) using the propensity score to estimate causal treatment effects in observational studies. Stat. Med. 2015;34(28):3661–3679.

17. Leyrat C, Seaman SR, White IR, et al. Propensity score analysis with partially observed covariates: How should multiple imputation be used? Stat. Methods Med. Res. 2019;28(1):3–19.

18. Penning de Vries BBL, Groenwold RH. A comparison of two approaches to implementing propensity score methods following multiple imputation. Epidemiol. Biostat. Public Health [electronic article]. 2017;14(4). (https://ebph.it/article/view/12630). (Accessed October 30, 2019)

19. R Core Team. R: A Language and Environment for Statistical Computing. Vienna, Austria: R Foundation for Statistical Computing; 2020.(http://www.R-project.org/)

20. Greifer N. WeightIt: Weighting for Covariate Balance in Observational Studies. 2020.(https://CRAN.R-project.org/package=WeightIt)

21. Greifer N. cobalt: Covariate Balance Tables and Plots. 2020.(https://CRAN.R-project.org/package=cobalt)

22. Canty A, Ripley B. boot: Bootstrap R (S-Plus) Functions. 2020.

23. Vaughan D, Dancho M. furrr: Apply Mapping Functions in Parallel using Futures. 2020.(https://CRAN.R-project.org/package=furrr)

24. Wickham H, Averick M, Bryan J, et al. Welcome to the Tidyverse. J. Open Source Softw. 2019;4(43):1686.

25. Harrell F. Regression Modeling Strategies: With Applications to Linear Models, Logistic and Ordinal Regression, and Survival Analysis. 2nd ed. Springer International Publishing; 2015 (Accessed February 15, 2019).(https://www.springer.com/la/book/9783319194240). (Accessed February 15, 2019)

26. Bahamyirou A, Blais L, Forget A, et al. Understanding and diagnosing the potential for bias when using machine learning methods with doubly robust causal estimators. Stat. Methods Med. Res. 2019;28(6):1637–1650.

27. Penning de Vries BBL, Smeden M van, Groenwold RHH. Propensity Score Estimation Using Classification and Regression Trees in the Presence of Missing Covariate Data. Epidemiol. Methods [electronic article]. 2018;7(1). (https://www.degruyter.com/view/j/em.2018.7.issue-1/em-2017-0020/em-2017-0020.xml). (Accessed December 4, 2019)

28. Austin PC. Variance estimation when using inverse probability of treatment weighting (IPTW) with survival analysis. Stat. Med. 2016;35(30):5642–5655.

29. Cole SR, Hernán MA. Constructing Inverse Probability Weights for Marginal Structural Models. Am. J. Epidemiol. 2008;168(6):656–664.

30. Austin PC, Stuart EA. The performance of inverse probability of treatment weighting and full matching on the propensity score in the presence of model misspecification when estimating the effect of treatment on survival outcomes. Stat. Methods Med. Res. [electronic article]. 2017; (https://journals.sagepub.com/doi/10.1177/0962280215584401). (Accessed March 8, 2020)

31. Bartlett JW, Hughes RA. Bootstrap inference for multiple imputation under uncongeniality and misspecification: Stat. Methods Med. Res. [electronic article]. 2020;(https://journals.sagepub.com/doi/10.1177/0962280220932189). (Accessed August 13, 2020)

32. Seaman SR, White I. Inverse Probability Weighting with Missing Predictors of Treatment Assignment or Missingness. Commun. Stat. - Theory Methods. 2014;43(16):3499–3515.

33. Qu Y, Lipkovich I. Propensity score estimation with missing values using a multiple imputation missingness pattern (MIMP) approach. Stat. Med. 2009;28(9):1402–1414.

34. Schomaker M, Heumann C. Bootstrap inference when using multiple imputation. Stat. Med. 2018;37(14):2252–2266.

35. von Hippel PT, Bartlett JW. Maximum likelihood multiple imputation: Faster imputations and consistent standard errors without posterior draws. ArXiv12100870 Stat [electronic article]. 2019; (http://arxiv.org/abs/1210.0870). (Accessed January 26, 2020)

