## Supplemental Table 1 for "Considerations for using multiple imputation in propensity score-weighted analysis"

**Supplemental Table 1: Predictor matrix**

| | | **Predictors** | | | | | | | | | | | | | | | | | | | | | | | |
|---|---|---|---|---|---|---|---|---|---|---|---|---|---|---|---|---|---|---|---|---|---|---|---|---|---|
| **Type** | **Response** | migr | age_log | age | age_sb | sex | sexFemale:age | who_sqrt | who | who_sb | who:sexFemale | who:age | viol | ses | ptsd | ptsd_sb | edu | child | smok | bp_log | bp | bp_sb | hgt_log | hgt | hgt_sb |
| b | *migr* | | | | | | | | | | | | | | | | | | | | | | | | |
| c | *age_log* | | | | | | | | | | | | | 1 | 1 | 1 | 1 | 1 | | | 1 | 1 | | | |
| pas | age | | 1 | | | | | | | | | | | | | | | | | | | | | | |
| pas | age_sb | | | 1 | | | | | | | | | | | | | | | | | | | | | |
| b | *sex* | | | | | | | | | | | | | | 1 | 1 | | | 1 | | 1 | 1 | | 1 | 1 |
| pas | sexFemale:age | | | 1 | | 1 | | | | | | | | | | | | | | | | | | | |
| c | *who_sqrt* | | | 1 | 1 | 1 | 1 | | | | | | 1 | 1 | 1 | 1 | | | | | | | | | |
| pas | who | | | | | | | 1 | | | | | | | | | | | | | | | | | |
| pas | who_sb | | | | | | | | 1 | | | | | | | | | | | | | | | | |
| pas | who:sexFemale | | | | | 1 | | | 1 | | | | | | | | | | | | | | | | |
| pas | who:age | | | 1 | | | | | 1 | | | | | | | | | | | | | | | | |
| b | *viol* | | | 1 | 1 | 1 | 1 | | 1 | 1 | 1 | | | | 1 | 1 | | | | | | | | | |
| o | *ses* | | | 1 | 1 | 1 | 1 | | 1 | 1 | 1 | | | | 1 | 1 | | 1 | | | | | | | |
| c | *ptsd* | | | 1 | 1 | 1 | 1 | | 1 | 1 | 1 | 1 | 1 | 1 | | | | | | | | | | | |
| pas | ptsd_sb | | | | | | | | | | | | | | 1 | | | | | | | | | | |
| o | *edu* | | | 1 | 1 | 1 | 1 | | | | | | | 1 | | | | | | | | | | | |
| o | *child* | 1 | | 1 | 1 | 1 | 1 | | | | | | | 1 | | | 1 | | | | 1 | 1 | | | |
| b | *smok* | 1 | | | | 1 | | | | 1 | 1 | | 1 | | | | | | | | 1 | 1 | | 1 | 1 |
| c | *bp_log* | | | 1 | 1 | 1 | 1 | | | | | | | | | | | | | | | | | | |
| pas | bp | | | | | | | | | | | | | | | | | | | 1 | | | | | |
| pas | bp_sb | | | | | | | | | | | | | | | | | | | | 1 | | | | |
| c | *hgt_log* | | | | | 1 | | | | | | | | | 1 | 1 | | | 1 | | 1 | 1 | | | |
| pas | hgt | | | | | | | | | | | | | | | | | | | | | | 1 | | |
| pas | hgt_sb | | | | | | | | | | | | | | | | | | | | | | | 1 | |

*The predictor matrix details how each partially observed variable was imputed. Horizontally the variables enter as the response, vertically the variables enter as the predictor when the corresponding cell is "1". The "type" column indicate how the variables should be modelled when entering as the response variable (b, binary; c, continuous; pas, passive; o, ordered/ordinal; u, unordered/nominal). Variables in red are part of the substantive model (the propensity score model); variables in italic are actively imputed. ":" denotes interaction between the variables on each side.*
